# Episodic memory deficits are associated with impaired insight in traumatic encephalopathy syndrome: Initial findings of the SNAP-CTE study

**DOI:** 10.1101/2021.04.20.21255372

**Authors:** Rowena E. A. Mobbs, Jennifer Batchelor, Alessandra K. Teunisse, Ellen Erskine, Eamon Brown, Joshua Hood, Anthony E. Mobbs, Clare L. Fraser, Alan J Pearce, Richard Stevenson, Roy G Beran, Anna Miskovic-Wheatley, Karina Chan, Alicia Ching Mun Foo, Reidar P. Lystad

## Abstract

Chronic traumatic encephalopathy (CTE) has been identified at post-mortem in Australian football codes players. Detailed and objective clinical and radiological characterization of patients at-risk of sporting and non-sporting repetitive concussive and subconcussive traumatic brain injury (RC/SCI) is important to our understanding of traumatic encephalopathy syndrome (TES) and CTE. This paper presents the initial findings of the symptomatology, neurocognitive, and pathophysiological changes in CTE (SNAP-CTE) study. A preliminary, retrospective, cohort study of 28 patients (25 males and 3 females) presenting with complaints of mood, behavioral, and cognitive decline, comprising TES, aged 24 to 78 years (*M* = 53, SD = 15.3) with at least 10 years of exposure to RC/SCI (mostly through contact sport) and cognitive decline were examined including: demographics; duration of play; age of first exposure to RC/SCI; and neuropsychology. Participants performed significantly worse in tests of auditory, visual, immediate, and delayed memory compared to a normative sample. Those with absent insight, compared to those with preserved insight, had a larger discrepancy between predicted and actual auditory memory scores, and had more severe mood disturbances. Insight, a hallmark sign of dementia, was either absent or impaired in a significant proportion (*M* = 36.4%, *95% CI* =17.2% - 59.3%) of the sample aged under 65, compared with normative epidemiological data. This indicates that RC/SCI may be associated with risk of an early onset dementia syndrome representing probably TES/CTE. CTE is increasingly of concern to the Australian community, and further research in this area is necessary. This retrospective preliminary study will form the basis of large, prospective, longitudinal cohort studies, assessing the natural history of TES/CTE in Australian athletes.

## 1 Introduction

Patients with traumatic encephalopathy syndrome (TES) and the pathological correlate, chronic traumatic encephalopathy (CTE), experience a neurodegenerative disorder (1). As with other types of dementia, in which diagnostic tools are not definitive (2), TES requires verification at autopsy to confirm the presence of CTE (2,3). The pathognomic neuropathological lesion is the accumulation of hyperphosphorylated Tau in neurons and astrocytes, in an irregular distribution around blood vessels at the depths of sulci in the neocortex (4). While CTE has been mostly reported in American football players (5), it has also been found in other sports including soccer (6), professional wrestling, hockey (3) and in Australian-based football (7,8). However, the incidence and prevalence of a TES, as per criteria (1), and the association between concussion/subconcussion burden and TES or CTE is not yet evidenced in Australia. During life, these athletes typically experience concussive and subconcussive head trauma across different sports or non-sporting circumstances, which may overlap.

TES has been characterized to include cumulative head trauma beyond two years; correlating to progressive and delayed subjective and objective features; support by emotional dysregulation; behavioral changes; and motor disturbance, that are otherwise unaccounted for (1). The consideration of this relatively new differential subtype of dementia has not formed part of a traditional neuropathological examination at autopsy (9) nor have typical panels of comorbid cerebral pathology across longitudinal studies (10,11).

Although there is a clear need for neuropathological biomarkers to identify CTE, what is also needed is a clearer neuropsychological profile of this dementia. There is a growing body of literature investigating whether multiple concussive or subconcussive events contribute to long-term neuropsychological impairment in athletes (12,13). Studies assessing the prevalence of cognitive impairment indicate that clinically diagnosed mild cognitive impairment (MCI) is more frequent in contact-sport athletes than would be expected, according to population estimates (14) or when compared to age-matched, non-contact, sport controls (15). Survey data, from retired American football players, indicates that concussion history is associated with both clinically diagnosed MCI and subjective memory impairments (16). Self- and spouse-reports of impairments, consistent with MCI, suggest MCI may occur at a higher prevalence in former contact-sport athletes than would be expected by population estimates (17). This means that former players of contact related sports are more likely to show signs of MCI, compared with the general population. Such athletes also perceive themselves as having greater executive dysfunction, when compared to normative data (18) and age-matched non-contact sport athletes (19). This suggests some insight into the symptoms experienced. The literature remains equivocal as to whether these subjective concerns reflect objective cognitive impairments that can be detected on formalized neuropsychological measures.

Episodic memory, considered the encoding, retention, and active retrieval of specific contextual information, is influenced by MCI (20) and intelligence quotient (IQ) (21,22). Memory is supported by three processes: encoding (the intake and organization of information); consolidation (the storage of information); and retrieval (accessing the information) (23). A prominent study of sporting related RC/SCI (n = 35), as compared with single-impact head trauma (n = 35) or healthy controls (n = 20), found that both injury groups showed significant consolidation and retrieval deficits, whereas the single-impact head trauma group showed greater difficulty encoding (23). Although, it is unclear which aspects of episodic memory deficits are related to CTE. Other studies have found that athletes with a history of concussion, compared to controls or athletes without concussion, perform worse on tasks of episodic memory (14,24–26). One study that compared retired football players with three or more concussions to those with fewer than three concussions found no difference in their performance on memory testing (20). However, the two groups did demonstrate different neural recruitment patterns, which suggested that the retired players with a history of three or more concussions may have functional inefficiencies in memory networks. Therefore, it is important to investigate all aspects of episodic memory including Auditory Memory Index, Visual Memory Index, Immediate Memory Index, and Delayed Memory Index in a cohort of retired athletes to determine which elements (namely encoding, consolidation, or retrieval) are impaired and how they differ from other types of dementia.

Many investigations have assessed the association between concussion history and cognitive decline to establish a dose-response frequency. It has been shown that reductions in executive functioning, episodic memory, and information processing occur in repetitive head trauma. Former athletes who sustained their last sports-related concussion in environments of potential subconcussive burden, over 30 years prior, have demonstrated significantly poorer performance on tasks of episodic memory and response inhibition (26) or significantly impaired performance in visuomotor reaction time, spatial working memory, associative learning, and intra-extra dimensional shift (27,28), compared to healthy former athletes with no history of concussion. More research is needed to determine if there is a dose-dependent effect of mild Traumatic Brain Injury (mTBI) and exposure to repetitive head trauma related to CTE.

As with other forms of dementia, impaired insight may be a key feature when considering a patient’s risk of TES/CTE. Loss of insight is a lack of concern or unawareness of the consequences of symptoms and occurs independently of other psychological mechanisms of the denial of symptoms (29). Although a loss of insight might be indicative of frontotemporal dementia (29), it has also been reported in people with CTE (3,30,31).It is important to determine the extent of loss of insight in people with CTE and how this affects the progression of TES. A discrepancy between carer and patient reported number, type, or severity of symptoms, suggestive of lack of insight by patients into their illness, can have diagnostic and therapeutic implications (32,33).

These findings commence the work of the symptomatology, neurocognitive, and pathophysiological changes in CTE (SNAP-CTE) study, which assesses in-life clinical, radiological, neurophysiological, and neuropsychological characteristics across the spectrum of sporting and non-sporting repetitive head trauma in the Australian population. The larger aim of this project is to identify future biomarkers of disease, and to understand neuropathological and neuropsychological correlates. The current paper aims to determine the baseline neuropsychological characteristics of both active and retired contact and collision sporting participants extending upon the work by Pearce et al. (27,28) which was limited to cognitive outcomes. The primary variable of interest was the difference between IQ predicted and actual scores on the WMS-IV. It was hypothesized that differences between predicted and actual scores, in favor of IQ, would be both significant and clinically unusual. An additional aim was to investigate whether the participants’ level of insight or alcohol use is associated with their cognitive ability and memory. It was hypothesized that insight, as obtained on collateral history from the main carer, is impaired in TES, and that insight is important in determining a neurodegenerative picture as part of a systematic approach to the detection and diagnosis of younger-and older-onset dementia (34,35). It was also hypothesized that more severe episodic memory impairment would be associated with reduced insight.

## 2 Method

### 2.1 Participants

Participants were recruited from those referred from the general neurology clinic, sports medicine clinic, geriatric clinic, neurosurgical clinic, neuro-ophthalmology or ophthalmology clinic, or in general practice who self-identified as at risk for CTE. Participants were seen by a neurologist and neuropsychologist/psychologist. TES was defined as at least one core feature of impairment in cognition, mood, behavior, and at least two supportive features of impulsivity, anxiety, apathy, suicidality, paranoia, headache, motor signs, decline, delayed onset according to the Montinegro criteria (36) in the absence of other clinical and radiological syndromes that might account for their presentation, such as that of Alzheimer’s disease, Lewy body dementia, frontotemporal dementia, Parkinsons plus disorders, and normopressure hydrocephalus.

The exclusion criteria were: aged less than 18 years; prior clinical diagnosis of radiologically apparent traumatic brain injury; previous neurosurgery, birth trauma, or serious malignancy; individuals who started engaging with collisions contact sport at an older age (>17 years old); and if participants were unable to complete the Wechsler scales. The final sample consisted of 28 people (25 male, 3 female) aged 24 to 78 (*M* = 53.0, SD = 15.3) who were identified as at risk of sequelae of repeated head trauma. Demographic information relating to the participants and sporting history associated with history of concussion is included in Table 1.

**Table 1.** Demographics and sport associated with concussion data (n=28)

| Characteristic | n (%) |
| --- | --- |
| Age Group |  |
| 24–34 Years | 2 (7.1) |
| 35–44 Years | 8 (28.6) |
| 45–54 Years | 6 (21.4) |
| 55–64 Years | 6 (21.4) |
| ≥ 65 Years | 6 (21.4) |
| Gender, n males | 25 (89.3) |
| Level of education |  |
| < 10 Years | 2 (7.1) |
| School Certificate | 8 (28.6) |
| Higher School Certificate | 6 (21.4) |
| 13–15 Years | 7 (25.0) |
| ≥ 15 Years | 5 (17.9) |
| Sport/Event associated with concussion history* |  |
| Rugby League/Union/AFL | 17 (60.7) |
| Football/Soccer | 3 (10.7) |
| Boxing | 2 (7.1) |
| Equestrian | 2 (7.1) |
| Mountain/BMX riding | 2 (7.1) |
| Other** | 2 (7.1) |
| Basketball | 1 (3.6) |
| Softball | 1 (3.6) |
| Ice Hockey | 1 (3.6) |
| Water Skiing | 1 (3.6) |
| Wrestling | 1 (3.6) |
| Judo | 1 (3.6) |
| Reported Concussions |  |
| ≤ 9 | 5 (17.9) |
| 10–19 | 8 (28.6) |
| 20–29 | 6 (21.4) |
| ≥ 30 | 9 (32.1) |

\*6 participants were associated with 2 or more different sports
\*\* Domestic violence and army involvement

### 2.2 Measures

Participants were assessed using the battery of subtests from the Wechsler Adult Intelligence Scale-Fourth Edition (WAIS-IV) and Wechsler Memory Scale-Fourth Edition (WMS-IV). There was some variety in the combination of subtests administered to different participants from these test batteries. Patients who completed face to face testing were administered the full battery (n = 22), while those assessed via telehealth completed fewer measures (n = 6), in some cases, only verbal or visual Wechsler subtest depending on the technology used. As a result, there was some variation in the number of indices that were able to be analyzed across participants. Tests from the Verbal Comprehension Index and Working Memory Index of the WAIS-IV (both n = 18) and the Auditory Memory Index of the WMS-IV (n = 17) were most frequently included in administration. Tests from the Visual Working Memory index of the WMS-IV were least frequently included in administration (n = 5). The proportion of full indices that were able to be included in the analysis are detailed in Table 2.

**Table 2.** Index Scores Compared to the norm (M = 100)

|  | Index | *n* | *M* | *SD* | *M diff.* | *t* | df | *p* |
| --- | --- | --- | --- | --- | --- | --- | --- | --- |
| WAIS-IV | VCI | 28 | 98.2 | 12.7 | -1.79 | -.743 | 27 | 0.464 |
|  | PRI | 16 | 102.7 | 14.5 | 2.75 | .758 | 15 | 0.460 |
|  | WMI | 28 | 99.8 | 15.7 | -.21 | -.072 | 27 | 0.943 |
|  | PSI | 13 | 101.7 | 12.7 | 1.69 | .482 | 12 | 0.639 |
|  | FSIQ | 12 | 103.8 | 14.4 | 3.83 | .922 | 11 | 0.376 |
|  | GAI | 16 | 98.6 | 20.0 | -1.44 | -.287 | 15 | 0.778 |
| WMS-IV | AMI | 28 | 87.9 | 18.8 | -12.07 | -3.391 | 27 | 0.002 |
|  | VMI | 12 | 92.1 | 15.9 | -7.92 | -1.726 | 11 | 0.112 |
|  | VWMI | 5 | 96.4 | 7.2 | -3.60 | -1.124 | 4 | 0.324 |
|  | IMI | 12 | 93.2 | 17.9 | -6.75 | -1.306 | 11 | 0.218 |
|  | DMI | 12 | 89.7 | 21.4 | -10.33 | -1.669 | 11 | 0.123 |

*Note.* VCI = Verbal Comprehension Index; PRI = Perceptual Reasoning Index; WMI = Working Memory Index; PSI = Processing Speed Index; FSIQ = Full Scale Intelligence Quotient; GAI = General Ability Index; AMI = Auditory Memory Index; VMI = Visual Memory Index; VWMI = Visual Working Memory Index; IMI = Immediate Memory Index; DMI = Delayed Memory Index.

Raw data, obtained from the WAIS-IV and WMS-IV, were analyzed using the Advanced Clinical Solutions (ACS) (37) software package to produce index scores for both batteries. ACS was also used to conduct predicted-difference discrepancy analyses of WMS-IV index scores, using Verbal Comprehension Index or General Ability Index as the predictor. Discrepancies which occurred in less than 10% of the normative sample were considered unusually large and clinically meaningful (38).

A hallmark sign of the progress of neurodegenerative dementia is insight being either impaired or absent (29). In this study, the history of symptoms, as reported by patient and their accompanying, main carer during neurological consultation, were considered to reflect a measure of insight. Partially reduced insight (namely impaired) was as defined by a disparity in the reported symptomatology between patient and carer, while absent insight was the belief from the patient that they had no neurological symptoms in contradiction to the carer, and confirmed on assessment. Emotional dysregulation characterized by irritability, easy anger, easy rage and pervasive low mood or anxiety, as well as cognitive decline, were qualitatively recorded.

### 2.3 Procedure

This study was approved by the human research ethics committee at Macquarie University (reference number: RM03100). All cognitive testing was conducted by clinical neuropsychological or post-graduate psychology students using standardized administration as per the test manuals via face-to-face assessment (n = 22) or via a telehealth program (n = 6) due to COVID-19 restrictions. Information about insight and alcohol use was recorded during consultation with a neurologist (RM), which in all cases preceded neuropsychological assessment. The examiners conducting the latter were blinded to information regarding insight and alcohol use.

### 2.4 Statistical Analysis

Using the data obtained from ACS, one-sample t-tests were conducted to compare the current sample to a normative sample for cognitive ability using SPSS software. For all analyses, a p-value <0.05 was considered significant. Descriptive statistics were used to outline potential significant traits of the cohort, such as reported number of concussions, education, and alcohol use. We compared the proportion of the cohort that was aged under 65 and showed signs of impaired or absent insight (which is consistent with dementia type disease) to the proportion of people living with dementia under the age of 65 as reported by the Australian Government (39). A non-parametric binomial test was used to calculate the 95% confidence interval to determine significance of incidence of early onset dementia in our sample defined as under the age of 65 compared with the general population under the age of 65.

Due to the small amount of research and data within the area of concussion and TES/CTE, confounding variables are presently difficult to control. Alcohol use was thought to be potentially a confounding variable for TES/CTE and, within this sample, there were approximately 43% of participants that either presently or historically had alcohol use disorder or engaged in binge drinking behavior. Most of the sample (57%) did not show signs of alcohol use disorder nor engaged in binge drinking behavior, and approximately 86% had no active alcohol use disorder or binge pattern. Due to the cohort size, any statistically significant relationship between alcohol use and increased predilection to concussion and TES/CTE cannot be reliably assessed. Trends within the descriptive statistics of this cohort can indicate potential traits of interest for further investigation within this cohort and were reported.

## 3 Results

Nearly all of the participants included in the study participated in sport (as can be seen in Table 1) with most engaging in a sport that would have involved contact or collisions between athletes or head impact to the ground (e.g., Rugby League/Union/Australian Football League, soccer, and boxing). The potential for whiplash as a form of repeated head trauma was noted. Participation in these environments was for a total period of greater than 10 years, with the years of potential exposure ranging from 11 to 50 years (average age of contact sport commencement was *M =* 8.2 years old *SD* = 3.5 year). Years of education completed ranged from 8 to 22 years (*M* = 12.7, *SD* = 3.1).

### 3.1 Comparing the cohort to the norm for cognitive ability

The scores obtained on the WAIS-IV and WMS-IV indices were compared to a normative sample (*M* = 100, *SD* = 15) using a one-sample t-test and these scores are presented in Table 2. The average score on the WAIS-IV Verbal Comprehension Index was 98 and on the Working Memory Index average was 100. There were no statistically significant differences found between participants’ performances on any measures of intelligence from the WAIS-IV compared to a normative sample. See Table 2 for the p-values. There was a statistically significant differences found between performances on one of the WMS-IV indices and the normative sample. Participants performed significantly worse on the Auditory Memory Index than the normative sample. There was no statistically significant difference between the groups’ performances on the Visual Working Memory index. Correlations between the subscales and full scales are presented in Table 3.

**Table 3.** Correlations Between VCI, PRI, WMI, PSI, FSIQ and GAI

|  |  | VCI | PRI | WMI | PSI | FSIQ | GAI |
| --- | --- | --- | --- | --- | --- | --- | --- |
| VCI | *r* |  |  |  |  |  |  |
|  | *p* | - |  |  |  |  |  |
|  | *n* |  |  |  |  |  |  |
| PRI | *r* | .473 |  |  |  |  |  |
|  | *p* | .064 | - |  |  |  |  |
|  | *n* | 16 |  |  |  |  |  |
| WMI | *r* | .397* | .626** |  |  |  |  |
|  | *p* | .037 | .010 | - |  |  |  |
|  | *n* | 28 | 16 |  |  |  |  |
| PSI | *r* | .602* | .517 | .424 |  |  |  |
|  | *p* | .029 | .085 | .149 | - |  |  |
|  | *n* | 13 | 12 | 13 |  |  |  |
| FSIQ | *r* | .873** | .782** | .697* | .784** |  |  |
|  | *p* | .000 | .003 | .012 | .003 | - |  |
|  | *n* | 12 | 12 | 12 | 12 |  |  |
| GAI | *r* | .725** | .806** | .633** | .629* | .941** |  |
|  | *p* | .001 | .000 | .009 | .028 | .000 | - |
|  | *n* | 16 | 16 | 16 | 12 | 12 |  |

*Note.* VCI = Verbal Comprehension Index; PRI = Perceptual Reasoning Index; WMI = Working Memory Index; PSI = Processing Speed Index; FSIQ = Full Scale Intelligence Quotient; GAI = General Ability Index
\*. Correlation significant at the 0.05 level (2-tailed).
\*\*. Correlation significant at the 0.01 level (2-tailed).

Discrepancy analyses were conducted using the predicted-difference method with Verbal Comprehension Index as the predictor variable for 10/18 (55.6%) of participants, and General Ability Index as the predictor variable for 8/18 (44.4%) of participants. Results of the analysis are presented in Table 4. There were statistically significant differences between predicted and obtained scores on the Auditory Memory Index, Visual Memory Index, Immediate Memory Index, and Delayed Memory Index, with obtained scores on all those indices being significantly lower than predicted scores. There was no statistically significant difference between predicted and obtained scores on the Visual Working Memory index.

**Table 4.** Predicted - Obtained Memory Index Scores

| Index | *n* | Predicted *M* (*SD*) | Observed *M* (*SD*) | Difference *M* (*SD*) | *t* value | *p* |
| --- | --- | --- | --- | --- | --- | --- |
| AMI | 28 | 99.9 (6.0) | 87.9 (18.8) | 12.0 (15.3) | 4.149 | <0.001 |
| VMI | 12 | 102.6 (7.9) | 92.1 (15.9) | 10.5 (10.2) | 3.574 | 0.004 |
| VWMI | 5 | 99.6 (5.3) | 96.4 (7.2) | 3.2 (5.4) | 1.324 | 0.256 |
| IMI | 12 | 102.8 (9.0) | 93.2 (17.9) | 9.59 (11.6) | 2.857 | 0.016 |
| DMI | 12 | 102.5 (8.1) | 89.7 (21.4) | 12.83 (14.7) | 3.031 | 0.011 |

*Note.* AMI = Auditory Memory Index; VMI = Visual Memory Index; VWMI = Visual Working Memory Index; IMI = Immediate Memory Index; DMI = Delayed Memory Index.

A clinically significant difference, at the individual level, was defined as a disparity between predicted and observed scores that occurs in <10% of the normative sample (40). The proportion of individuals obtaining clinically significant differences across the WMS-IV indices are presented in

Table 5 for each of three cut-off values (41). The highest proportion of clinically significant cases was found on the Immediate Memory Index, with 41.7% of individuals obtaining a difference that occurs in less than 10% of the population.

**Table 5.**
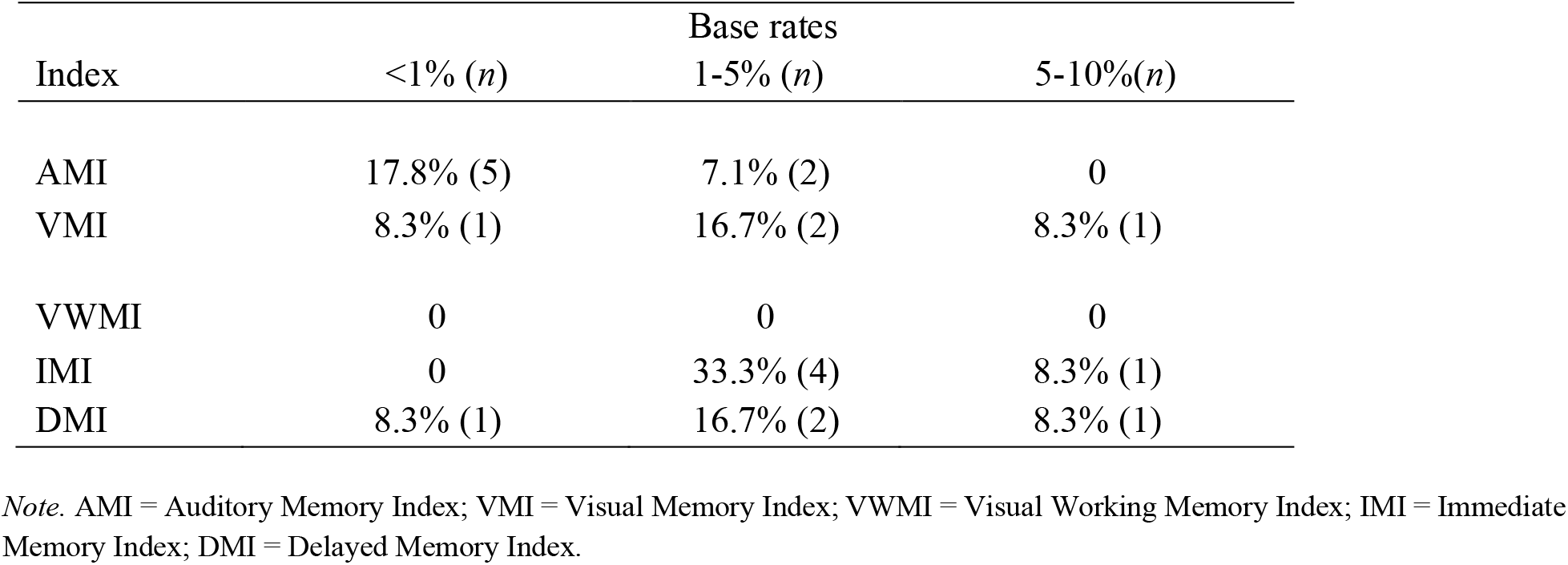
Proportions of Cases Meeting Clinical Significance at Each of Three Base-Rate Categories for Predicted-Obtained Discrepancy Scores

### 3.2 Insight capacity

Out of the 28 participants, 64.3% (n=18) demonstrated preserved insight, while 35.7% participants had either impaired (n=6) or absent (n=4) insight. See

Table 6 for statistics of the cohort partitioned by level of insight. There appeared to be no significant difference in age at presentation (*M*, absent/impaired = 52 years, preserved = 53 years), age at commencement of play (*M*, absent/impaired = 10 years, preserved = 8 years) or years of education (*M*, absent/impaired = 12 years, preserved = 13 years). There did appear to be a difference between years of exposure, with the absent/impaired cohort (*M* = 19 years) showing fewer total years of exposure compared with the preserved cohort (*M* = 25 years).

**Table 6.** Cohort characteristics by level of insight

|  | Absent/Impaired<br>(n=10) | Preserved<br>(n=18) | Odds Ratio<br>(95%CI) | p-value |
| --- | --- | --- | --- | --- |
| Age at presentation | 52.7 (15.7) | 53.2 (15.5) | 1.00 (0.95, 1.06) | 0.937 |
| Education (Years) | 12.3 (3.7) | 12.9 (2.8) | 1.07 (0.83, 1.44) | 0.595 |
| Age of Commencement | 9.5 (3.5) | 7.5 (3.4) | 0.84 (0.65, 1.06) | 0.154 |
| Total years exposure<br>contact | 19.2 (4.3) | 24.9 (9.8) | 1.12 (1.00, 1.33) | 0.113 |
| Concussions | 38.1 (36.4) | 27.8 (31.4) | 0.99 (0.97, 1.01) | 0.425 |
| Verbal Comprehension<br>Index | 92.5 (12.6) | 101.4 (11.9) | 1.07 (1.00, 1.16) | 0.088 |
| Working Memory Index | 100.7 (17.2) | 99.3 (15.4) | 0.99 (0.94, 1.05) | 0.815 |
| Auditory Memory Index | 83.8 (17.9) | 90.2 (19.5) | 1.02 (0.98, 1.07) | 0.384 |
| Duration mood at<br>presentation (years) | 9.0 (9.6) | 4.2 (7.6) | 0.94 (0.84, 1.03) | 0.170 |
| Duration cognitive decline<br>at presentation (years) | 6.4 (7.2) | 2.9 (3.9) | 0.88 (0.69, 1.02) | 0.161 |
| ACE-III / ACE-R at<br>presentation /100<sup>1</sup> | 87.4 (13.2) | 92.7 (4.4) | 1.09 (0.97, 1.31) | 0.254 |

<sup>1</sup> ACE data missing for n=2 patients with absent/impaired insight and n=5 patients with preserved insight.

According to the Australian Institute of Health and Welfare, in 2019, approximately 0.13% (n=26,871) of Australians under 65 years were living with dementia (39,42). Within this study’s cohort who reported to a neurologist, 78.5% (n = 22, *M* = 47.1, S.D. = 11.2) were under the age of 65. Within this subset, it was observed that 36.4% (n=8) of the cohort had either impaired or absent insight. The binomial test indicated that the proportion of insight impairment in our cohort was significantly higher than the prevalence of dementia among Australian aged under 65 years (n=22, k=8, p_k_=0.13%, p < 0.001).

There were variations in the number of concussions sustained between the absent/impaired (*M* = 38.1) and preserved (*M* = 27.8) insight. But when examining the median number of concussions, due to the right-skew of the data, this difference reduces (absent/impaired Median = 20, preserved Median = 18.5). Due to the small sample size and the skew of the data it was unclear if there is a relationship just based on group averages. Working Memory Index did not appear to vary with insight (*M, absent/impaired* = 100.7 and *M, preserved* = 99.3). The Verbal Comprehension Index scores did appear to differ, where the absent/impaired group had lower average scores (*M* = 92.5) compared with the preserved group (*M* =101.4) but this difference failed to reach significance. The absent/impaired group had lower Auditory Memory Index values (*M* = 83.8) compared with the preserved group (*M* = 90.2). This suggests that there could be a relationship between Auditory Memory Index and insight, whereby if auditory memory is impacted, so too will be insight.

Although not all groups had complete ACE-III (Addenbrooke’s Cognitive Examinations) scores (43), the absent/impaired insight group had lower scores (*M* = 87.4) compared the preserved insight capacity group (*M* = 92.7). The duration of reported mood disturbance at presentation and duration of reported cognitive decline at presentation, was on average considerably higher in the absent/impaired group (cognition *M* = 6.4, mood *M* = 9.0) compared with the preserved group (cognition *M* = 2.9, mood *M* = 4.2). This showed that where the insight of participants was altered, participants were experiencing cognition and mood changes for twice as long before presentation.

### 3.3 Alcohol use and cognitive ability

Of the participants who completed the ACE-III scores (*n* = 21), the total average was low, being approximately 1.6 points higher than what is considered clinically significant for dementia (*M* = 90.6, *SD* = 8.9). See Table 7 for the ACE-III scores presented by group. It appeared that the non-drinking group out-performed their drinking counterparts, by showing ACE-III scores relatively higher than what was considered clinically significant for dementia (*M* = 92.2, SD = 4.3) compared with the drinking group (*M* = 88.1, SD = 13.5). Table 7 also presents ACE-III scores subdivided into level of insight. The average ACE-III scores did not appear to differ between groups based on drinking status and level of insight except for those participants in the alcohol use disorder/binge drinking group who also had absent/impaired insight. Their average ACE-III scores were almost 20 points lower. However, there were only 2 participants in this group, suggesting that this may be a statistical artefact.

**Table 7.**
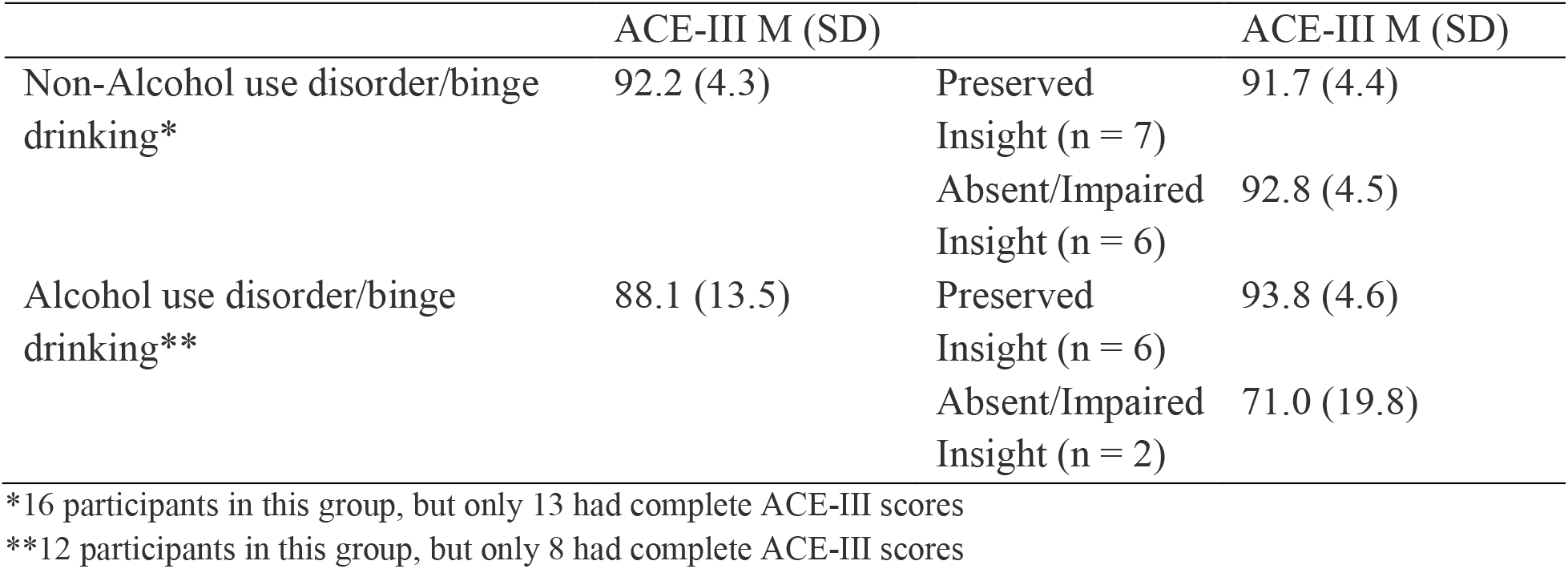
ACE-III scores presented by group

## 4 Discussion

This was the first Australian study to assess the full spectrum of neuropsychological changes in a group of RC/SCI patients who met criteria for TES compared to normative data. There was support for the hypothesis, such that there were differences between the participants predicted IQ scores and actual IQ scores in various facets of episodic memory. Participants in this cohort performed significantly worse on measures of auditory, visual, immediate, and delayed memory compared to a normative sample. Previous alcohol use disorders may exacerbate disease progression. Participants with impaired or absent insight had poorer verbal comprehension compared with the preserved insight group. Participants with absent or impaired insight had a larger discrepancy between predicted and actual scores on measures of auditory memory, compared to the other two groups. Mood disturbances were higher in the absent/impaired insight group compared to the preserved insight group. Participants with absent or impaired insight had less years of exposure but many more reported concussions, however the skew of the data meant that when examining the median number of reported concussions, this difference reduced. 78.5% of the sample were under the age of 65 yet reporting to a neurologist with concerns about their cognitive functioning. This is in stark contrast to the 0.1% of the Australian population who are currently suffering dementia and are under the age of 65 (39,42) suggesting that RC/SCI is associated with early onset dementia.

Although the severity of the behavioral and psychological disturbance was assessed qualitatively, rather than quantitatively, the cohort were consistently described to have impairment at first neurological consultation. Examples include: paranoid ideation, tendency of anger or rage, and cognitive impairment. Carers consistently reported troubles with irritability, social demeanor, anger, and physical rage accompanying memory impairment that were consistent with neuropsychological findings. These behavioural and psychological disturbances have been similarly described in the review by Montenigro et al. (36). Also, it is likely that alcohol use may be contributing to the neurodegenerative processes of RC/SCI, yet it is unclear if alcohol use exacerbates the progression of disease or if alcohol is a confounding variable. Further investigation is required to determine the true impact of alcohol use as related to RC/SCI and TES.

The value of this study is that it integrated measurement of intellect, working memory, and episodic memory. The study considered performance on tests of working memory and episodic memory relative to norms, rather than to a control group, which could have inherently differed in terms of intellect and by extension, memory. Performance on measures of encoding and retention were impaired relative to predictions derived from IQ. Performance on tests of working memory, which represents one aspect of attention, was not impaired. These findings extend recent reports (23,44) of an impairment of episodic memory in this population and contribute to the literature by demonstrating a significant disparity between the individual’s IQ and memory (45). These findings cannot be explained by reference to poorly matched groups or other methodological limitations. The findings add to the growing body of literature indicating that this group experiences measurable and significant cognitive impairment.

While this study had a small sample, the cohort is suggestive of neurocognitive issues occurring much earlier than anticipated with a traditional dementia onset – including younger onset dementia (34). There was a 12-point difference, on average, in this group between the predicted Auditory Memory Index and the actual result. There appeared to be a trend that number of years of education, may have a protective effect on Auditory Memory Index scores. Given the small sample size, this should be investigated further.

There seemed to be no clear available demographic factors amongst the cohort that increased their likelihood of sustaining a concussion, except for a small positive trend regarding years of play. This relationship between has also been found by Rafferty et al. (46) who noted that playing more than 25 matches increased the likelihood of receiving a concussion. It did appear that alcohol use disorder, alcohol use, and binge drinking, whether concurrent or historic did have either an exacerbating or confounding role within concussion/subconcussion and TES. Further investigation is required to determine the true nature of alcohol use within the disease progression of TES due to exposure to concussive events. Alcohol use disorder or binge use may be reactive to the emergence of other psychological or cognitive symptoms, consistent with described ‘self-medication’ by some within our cohort. It may also be secondary to behavioral dysregulation as a result of cognitive impairment and TES, or contribute to further cognitive behavioral impairment. Alcohol use is now interpreted not only as a cause of TBI but a secondary disorder after TBI, as well as a predictive factor for outcome (47). Although no patients were diagnosed, those with active alcohol use disorders were provided prophylactic thiamine towards the prevention of Wernicke-Korsakoff syndrome. An alcohol-related dementia cannot be excluded as contributing to cognitive impairment, however the presence of comparable impairment across those with or without a background alcohol history in the current cohort would favor RC/SCI as a causal factor.

One of the central limitations of the study is the self-selection bias of individuals who volunteered to participate in this behavioral research and the typical characteristics of participants such as greater years of education and higher IQ scores (48,49). Further, it is likely that only those concerned with their cognitive ability are seeking treatment from a neurologist. This self-selection bias may have affected the results. The social desirability of participating in a research study for many athletes often typically derives from a strong personal interest in the results of the study (50), with many acknowledging a concomitant desire to contribute to a body of knowledge that will contribute to both awareness and intervention/prevention of the issue under investigation. As a result, athletes presenting to behavioral research may not be representative of the general sporting population. Additionally, the number of concussive episodes in the sample was largely based on the experience of the patient and their family and is subject to include recall bias. Further investigation would be needed to exclude recall bias and other noted confounding variables. Concussive events in contact sport have historically been considered a “normal” aspect of play, being largely under-evaluated and under-declared (51). This may bias the retrospective recollection of such incidents and some studies have not investigated history of concussion as a predictor variable as they could not confidently ascertain the number of concussions experienced by each athlete (15).Cognitive insight was measured subjectively by both the participant and their carer/spouse. Future research should employ a more objective measure of cognitive insight. This study was underpowered and as a result, we could only report on trends in the data and not control for confounding variables, such as alcohol use.

### 4.1 Conclusion

Within this small retrospective cohort, the criteria for TES were fulfilled and a significant reduction in episodic memory was identified, with exclusion on clinic/radiological assessment of other causes. This study confirms 95% confidence that the true population proportion of diminished insight capacity (one of the hallmark signs of dementia), within a group of post-concussed or subconcussive individuals aged under 65 that reported to a neurologist falls between 17.2% and 59.3%, which is significantly higher than the rate of dementia observed within the general population aged under 65 within Australia. The research team acknowledges the limitations of most retrospective studies, whereby the significant findings may not account for other co-related confounding variables. The aim is to conduct further studies to understand the true nature of these relationships. This means that it is likely that concussive and subconcussive events may increase the risk of early onset dementia. The plan is to develop suitably powered large cohort studies assessing the natural history of TES/CTE for the population from the SNAP-CTE study.

## Data Availability

Data is available upon request

## 5 Author Contributions

Dr Mobbs had full access to all the data in the study and takes responsibility for the integrity of the data and the accuracy of the data analysis.

Study concept and design: All authors.

Acquisition, analysis, or interpretation of data: All authors.

Drafting of the manuscript: All authors.

Critical revision of the manuscript for important intellectual content: All authors.

Administrative, technical, or material support: Teunisse

Study supervision: Mobbs, Batchelor

